# Multimorbidity and blood pressure control: a cross-sectional analysis among 67,385 adults with hypertension in Canada

**DOI:** 10.1101/2023.07.24.23293126

**Authors:** Tu N Nguyen, Sumeet Kalia, Peter Hanlon, Bhautesh D Jani, Barbara I Nicholl, Chelsea D. Christie, Babak Aliarzadeh, Rahim Moineddin, Christopher Harrison, Clara Chow, Martin Fortin, Frances S Mair, Michelle Greiver

**Affiliations:** Westmead Applied Research Centre, Sydney Medical School, Faculty of Medicine and Health, University of Sydney, Sydney, New South Wales, Australia; Department of Family and Community Medicine, Temerty Faculty of Medicine, University of Toronto, Toronto, ON, Canada; General Practice and Primary Care, School of Health and Wellbeing, University of Glasgow, UK; Menzies Centre for Health Policy and Economics, School of Public Health, Faculty of Medicine and Health, University of Sydney, Sydney, New South Wales, Australia; Department of family medicine and emergency medicine, Université de Sherbrooke, Saguenay, QC, Canada; Department of Family and Community Medicine, North York General Hospital, Toronto, ON, Canada

**Keywords:** hypertension, high blood pressure, blood pressure control, multimorbidity, primary care

## Abstract

**Background:** There has been conflicting evidence on the association between multimorbidity and blood pressure (BP) control. This study aimed to investigate this associations in people with hypertension attending primary care in Canada, and to assess whether individual long-term conditions are associated with BP control.

**Methods:** A cross-sectional study in people with hypertension attending primary care in Toronto between January 01 2017 and December 31 2019. Uncontrolled BP was defined as systolic BP≥140 mmHg or diastolic BP≥90 mmHg. A list of 11 a priori selected chronic conditions was used to define multimorbidity. Multimorbidity was defined as having ≥1 long-term condition in addition to hypertension. Logistic regression models were used to estimate the association between multimorbidity (or individual long-term conditions) with uncontrolled BP.

**Results:** A total of 67,385 patients with hypertension were included. They had a mean age of 70, 53.1% were female, 80.6% had multimorbidity, and 35.7% had uncontrolled BP. Patients with multimorbidity had lower odds of uncontrolled BP than those without multimorbidity (adjusted OR 0.72, 95% CI 0.68-0.76). Among the long-term conditions, diabetes (aOR 0.73, 95%CI 0.70-0.77), heart failure (aOR 0.81, 95%CI 0.73-0.91), ischemic heart disease (aOR 0.74, 95%CI 0.69-0.79), schizophrenia (aOR 0.79, 95%CI 0.65-0.97), depression/anxiety (aOR 0.91, 95%CI 0.86-0.95), dementia (aOR 0.87, 95%CI 0.80-0.95), and osteoarthritis (aOR 0.89, 95%CI 0.85-0.93) were associated with a lower likelihood of uncontrolled BP.

**Conclusion:** We found that multimorbidity was associated with better BP control. Several conditions were associated with better control, including diabetes, heart failure, ischemic heart disease, schizophrenia, depression/anxiety, dementia, and osteoarthritis.

## Introduction

Hypertension is a risk factor for cardiovascular disease and other chronic health conditions. The prevalence of hypertension increases with age, with a prevalence of only 27% in people younger than 60 years, but 74% in people aged 80 and older.^1^ Globally, blood pressure (BP) control is still suboptimal, with control rates of only approximately 20% for people with hypertension.^2^ Poor control of hypertension can increase the risk of heart failure, coronary heart disease, peripheral artery disease, renal failure, stroke and dementia, and therefore, increase the risk of developing multimorbidity in people with hypertension.^3^ Multimorbidity, defined as having two or more chronic conditions, has become a primary healthcare concern.^4, 5^

Two out of three people with hypertension have additional long-term condition.^6^ A systematic review of 45 studies conducted from 2007 to 2017 showed that the overall prevalence of multimorbidity was 66.1% (when multimorbidity was defined as having ≥ 2 chronic conditions), and 44.2% (when multimorbidity was defined as having ≥ 3 chronic conditions) in older adults in high-income countries.^7^ In the United States, 81 million adults were estimated to have multimorbidity in 2020.^8^ In Canada, the prevalence of multimorbidity is also high, with a reported prevalence ranging from around 30% to 70% in primary care settings, and around 17% to 59% in the general population.^9^ Multimorbidity increases healthcare costs and healthcare utilisation including hospitalisations.^10, 11^

Several studies have examined the impact of multimorbidity on BP control, and there has been conflicting evidence on the association between multimorbidity and hypertension control.^12–17^ While some studies show that the presence of multimorbidity was positively associated with BP control,^12, 16, 18^ other studies suggested that people with more comorbidities had poorer management and control of hypertension.^15, 17^ There is minimal evidence of this association in the Canadian population. Therefore, in this study, we aimed to investigate the association between multimorbidity and BP control in patients attending primary care in Canada, and to assess if individual comorbidities are associated with BP control.

## Materials and Methods

### Data source

This study used data from the University of Toronto Practice-Based Research Network (UTOPIAN), a primary care electronic medical records (EMRs) database. The UTOPIAN database contains de-identified records from participating family medicine clinics in Ontario, Canada, with most providers practicing in the Greater Toronto Area.^19^ Data include de-identified patient-level information on various factors including demographics, medical diagnoses, procedures, medications, immunizations, laboratory test results, vital signs, risk factors, and clinical notes.

### Study design and eligible patients

This is a cross-sectional study using the UTOPIAN 2021Q4 database with a start date of January 01 2017 and an end date of December 31 2019. This end date was chosen to avoid the COVID-19 pandemic-related effects on the data. The data were accessed for research purposes from June 17 2022 to December 16 2022. We identified a cohort of patients who met the following selection criteria; had hypertension before December 31 2019; had blood pressure recorded between Jan 01 2017 and Dec 31 2019; and were at least 45 years old when their blood pressure was measured. **Figure 1** provides a flowchart for the cross-sectional cohort.

**Figure 1:**
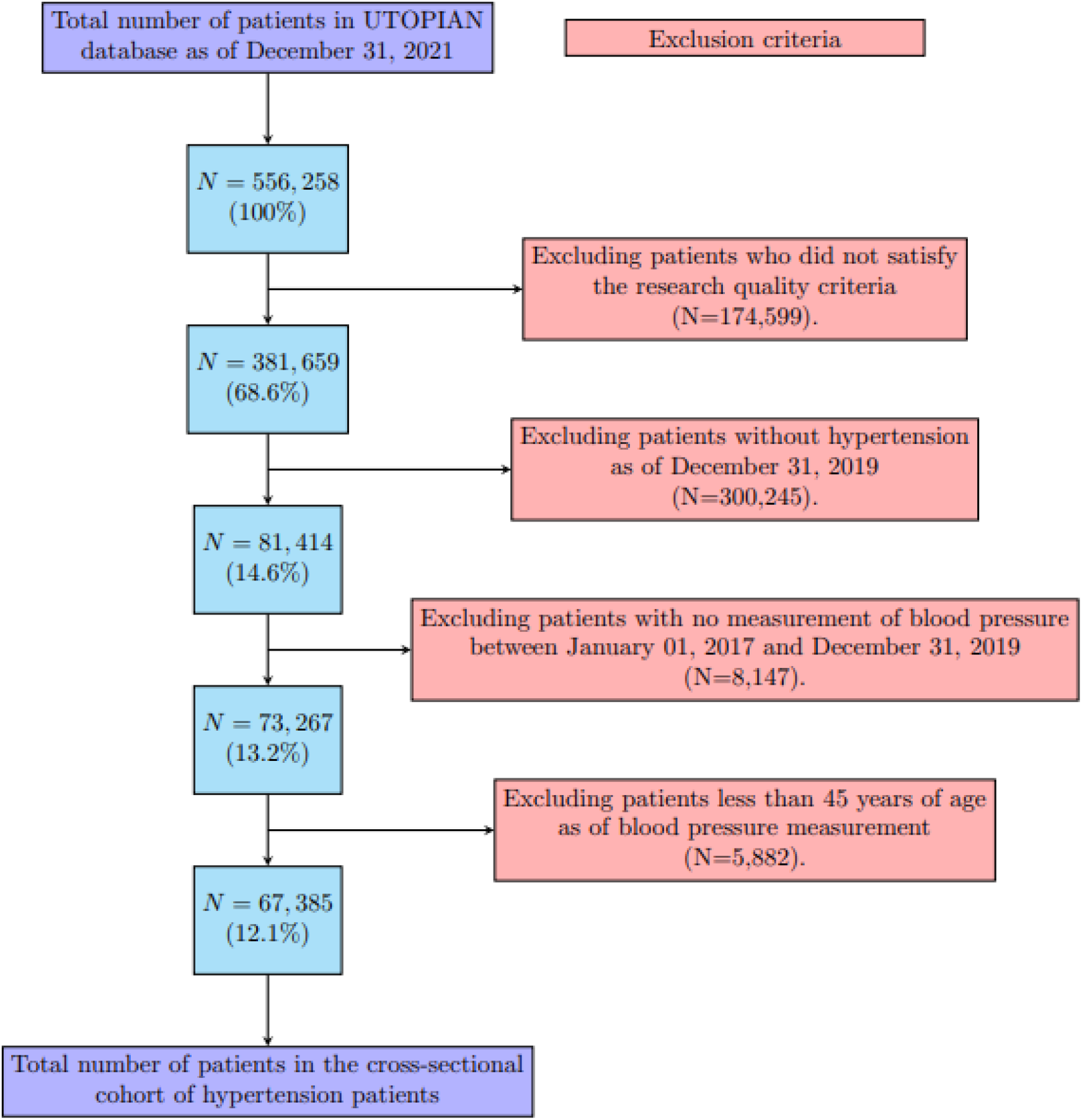
Flowchart for the generation of this cross-sectional cohort

We defined the hypertension phenotype using the following criteria (Appendix 2):

1. Free text documentation of hypertension was found within the (past or present) health condition section of the cumulative patient profile, including the following terms: hypertension, hypertensive, htn; OR
2. Anti-hypertensive medication was prescribed and an elevated blood pressure reading was recorded at any point in the EMR (elevated blood pressure reading is defined as systolic blood pressure ≥ 140 mmHg or diastolic blood pressure ≥ 90 mmHg); OR
3. Anti-hypertensive medication was prescribed and a billing record with the diagnosis code for hypertension (ICD-9 code 401) was recorded at any point in the EMR; OR
4. A billing record with the diagnosis code for hypertension (ICD-9 code 401) and an elevated blood pressure reading was recorded at any point in the EMR (elevated blood pressure reading is defined as systolic blood pressure ≥ 140 mmHg or diastolic blood pressure ≥ 90 mmHg)

### Multimorbidity

Multimorbidity was defined as having one or more long-term conditions in addition to hypertension. This definition of multimorbidity is similar to definitions used in other studies (REFs).^20–22^ A list of 11 a priori selected chronic conditions was used to define multimorbidity, in alignment with the Centers for Disease Control and Prevention (CDC) recommendations for defining and measuring multiple chronic conditions when using Medicare claims.^23^ These long-term condition are: ischemic heart disease, heart failure, atrial fibrillation, diabetes, chronic obstructive pulmonary disease, asthma, chronic kidney disease (chronic kidney disease stage 3 or more, or dependence on transplant or dialysis), cancers (including breast cancer, cervical cancer, lung cancer, colorectal cancer, ovarian cancer, pancreatic cancer, prostate cancer, thyroid cancer), osteoarthritis, dementia, depression and/or anxiety, and schizophrenia.

### Covariate definitions

We used the most recent information on Body Mass Index (BMI) and smoking status as of December 31, 2019. BMI was grouped into underweight (≤18.4), normal (18.5-24.9), overweight (25.0-29.9), obese class I (30.0-34.9), obese class II (35.0-39.9), and obese class III (≥40.0). We calculated age (in years) on the date when the blood pressure measurement was recorded between January 01, 2017 and December 31, 2019. We used rurality and neighbourhood income quintiles (as an indicator of socioeconomic status) using data from the 2016 Statistics Canada Census.^24^

We identified different classes of hypertension medication (including angiotensin converting enzyme inhibitors (ACEi), angiotensin II receptor blocker (ARBi), calcium channels, beta-blockers, diuretics, and other classes of hypertension medication) that were prescribed during the six months prior to the blood pressure measurement. The full description of drug names for each class of hypertension medication is provided in Appendix section (i), along with the algorithm used to determine the presence of hypertension in EMRs (see Appendix section (ii)).

### Outcome measure

The primary outcome measurement was the most recent BP measure recorded during the study period (January 01, 2017 to December 31, 2019). Uncontrolled BP was defined as having either systolic blood pressure ≥140 mmHg or diastolic blood pressure ≥90 mmHg for the last BP measurement prior to December 31 2019. The BP treatment target was chosen to align with the Hypertension Canada’s 2016 Canadian hypertension education program guidelines for blood pressure measurement, diagnosis, assessment of risk, prevention, and treatment of hypertension that reflected the practice during the study period .^25^ The SBP treatment goal is a pressure level of < 140 mm Hg (Grade C). The DBP treatment goal is a pressure level of < 90 mm Hg (Grade A). These targets were established using Office-based BP Measurements (OBPM).^25^ For patients with multiple BP readings recorded on the same day at the same visit (e.g. using an automated BPtru machine), we took the average measurement of the systolic and diastolic blood pressure measures on that day.

We also conducted a sensitivity analysis, with a BP target of < 130/80 mmHg, as recommended by the 2018 ESC/ESH Guidelines for the management of arterial hypertension^26^, and the 2017 ACC/AHA/AAPA/ABC/ACPM/AGS/APhA/ASH/ASPC/NMA/PCNA Guideline for the Prevention, Detection, Evaluation, and Management of High Blood Pressure in Adults.^27^

### Statistical analysis

We used descriptive statistics to summarize patient demographics and clinical characteristics associated with controlled and uncontrolled blood pressure. We used means, standard deviations, and median, minimum and maximum to summarize continuous variables, while frequencies and percentages were used to summarize categorical variables. To reduce the risk of re-identification, our reports exclude any cell count of five or fewer. Comparisons between patients with controlled and uncontrolled BP were assessed using Pearson chi-square tests for categorical covariates and using two-sample t-tests for continuous covariates.

We fitted logistic regression models using the Generalized Estimating Equations (GEE) with an exchangeable working correlation matrix to assess the association between multimorbidity (or individual long-term condition) on uncontrolled BP. We reported both the unadjusted odds ratios and adjusted odds ratios. To account for multicollinearity, we fitted separate logistic models using GEE for multimorbidity and different comorbidities, respectively. We a priori specified age, sex, income quintiles, smoking BMI as potential confounders (i.e. common cause) for multimorbidity and for the outcome of BP control, while different classes of hypertension medications were considered potential mediators on the pathway between multimorbidity and BP control (see Supplementary Figure S1). In GEE models, we adjusted for the following covariates: age, sex, income quintiles, smoking status, BMI, number of primary care visits, along with the multimorbidity status.

In a sensitivity analysis, we used a reduced threshold for blood pressure control (<130/80 mm Hg) and excluded BMI and smoking status in the regression models to include patients that did not have BMI or smoking status recorded.

### Ethics approval

This study was approved by the Research Ethics Board at the University of Toronto (protocol number 00043354).

## Results

### Sample characteristics

A total of 67,385 patients who satisfied the selection criteria were included in this study. Patients in the sample had a mean age of 70.1 years (SD 11.8) and 53.1% were female. The prevalence of overweight and obesity was 66%, with a greater proportion of patients with class II and class III obesity in the uncontrolled BP group compared to the controlled BP group. There was no difference in socioeconomic status between the two groups. (**Table 1**)

**Table 1:** Patient characteristics.

|  | **Controlled BP**<br>**(N=43303)** | **Uncontrolled**<br>**BP**<br>**(N=24082)** | **Overall**<br>**(N=67385)** |
| --- | --- | --- | --- |
| **Age (years)** |  |  |  |
| Mean (SD) | 70.3 (11.6) | 69.6 (12.2) | 70.1 (11.8) |
| Median [Min, Max] | 70.0 [47.0, 111] | 69.0 [47.0, 110] | 70.0 [47.0, 111] |
| **Age group (years)** |  |  |  |
| 45-74 years | 30562 (70.6%) | 17256 (71.7%) | 47818 (71.0%) |
| 75-84 years | 8927 (20.6%) | 4486 (18.6%) | 13413 (19.9%) |
| 85+ years | 3814 (8.8%) | 2340 (9.7%) | 6154 (9.1%) |
| **Sex** |  |  |  |
| Female | 22637 (52.3%) | 13132 (54.5%) | 35769 (53.1%) |
| Male | 20666 (47.7%) | 10950 (45.5%) | 31616 (46.9%) |
| **Income Quintiles** |  |  |  |
| 1 (low income) | 8984 (20.7%) | 5090 (21.1%) | 14074 (20.9%) |
| 2 | 7812 (18.0%) | 4328 (18.0%) | 12140 (18.0%) |
| 3 | 7264 (16.8%) | 3938 (16.4%) | 11202 (16.6%) |
| 4 | 7572 (17.5%) | 4177 (17.3%) | 11749 (17.4%) |
| 5 (high income) | 10693 (24.7%) | 5976 (24.8%) | 16669 (24.7%) |
| Missing | 978 (2.3%) | 573 (2.4%) | 1551 (2.3%) |
| **Multimorbidity** |  |  |  |
| No | 7473 (17.3%) | 5574 (23.1%) | 13047 (19.4%) |
| Yes | 35830 (82.7%) | 18508 (76.9%) | 54338 (80.6%) |
| **Body mass index level** |  |  |  |
| Missing | 7831 (18.1%) | 5018 (20.8%) | 12849 (19.1%) |
| 18.4 or less (Underweight) | 267 (0.6%) | 145 (0.6%) | 412 (0.6%) |
| 18.5 - 24.9 (Normal) | 6453 (14.9%) | 3220 (13.4%) | 9673 (14.4%) |
| 25 - 29.9 (Overweight) | 13470 (31.1%) | 6999 (29.1%) | 20469 (30.4%) |
| 30 - 34.9 (Obese Class I) | 9278 (21.4%) | 5095 (21.2%) | 14373 (21.3%) |
| 35 - 39.9 (Obese Class II) | 3782 (8.7%) | 2208 (9.2%) | 5990 (8.9%) |
| 40 or more (Obese Class III) | 2222 (5.1%) | 1397 (5.8%) | 3619 (5.4%) |
| **Most recent smoking status** |  |  |  |
| Missing | 11062 (25.5%) | 6740 (28.0%) | 17802 (26.4%) |
| Current | 4219 (9.7%) | 2542 (10.6%) | 6761 (10.0%) |
| Non-Smoker | 19999 (46.2%) | 10868 (45.1%) | 30867 (45.8%) |
| Past | 8023 (18.5%) | 3932 (16.3%) | 11955 (17.7%) |
| **Cancer** |  |  |  |
| No | 37881 (87.5%) | 21349 (88.7%) | 59230 (87.9%) |
| Yes | 5422 (12.5%) | 2733 (11.3%) | 8155 (12.1%) |
| **Ischemic Heart disease** |  |  |  |
| No | 37182 (85.9%) | 21679 (90.0%) | 58861 (87.4%) |
| Yes | 6121 (14.1%) | 2403 (10.0%) | 8524 (12.6%) |
| **Heart failure** |  |  |  |

|  | **Controlled BP**<br>**(N=43303)** | **Uncontrolled BP**<br>**(N=24082)** | **Overall**<br>**(N=67385)** |
| --- | --- | --- | --- |
| No | 40754 (94.1%) | 23082 (95.8%) | 63836 (94.7%) |
| Yes | 2549 (5.9%) | 1000 (4.2%) | 3549 (5.3%) |
| **Atrial fibrillation** |  |  |  |
| No | 38362 (88.6%) | 21878 (90.8%) | 60240 (89.4%) |
| Yes | 4941 (11.4%) | 2204 (9.2%) | 7145 (10.6%) |
| **Diabetes Mellitus** |  |  |  |
| No | 28126 (65.0%) | 17210 (71.5%) | 45336 (67.3%) |
| Yes | 15177 (35.0%) | 6872 (28.5%) | 22049 (32.7%) |
| **Chronic Kidney disease** |  |  |  |
| No | 35292 (81.5%) | 20021 (83.1%) | 55313 (82.1%) |
| Yes | 8011 (18.5%) | 4061 (16.9%) | 12072 (17.9%) |
| **Chronic Obstructive Pulmonary Disease** |  |  |  |
| No | 39365 (90.9%) | 22197 (92.2%) | 61562 (91.4%) |
| Yes | 3938 (9.1%) | 1885 (7.8%) | 5823 (8.6%) |
| **Osteoarthritis** |  |  |  |
| No | 26654 (61.6%) | 15500 (64.4%) | 42154 (62.6%) |
| Yes | 16649 (38.4%) | 8582 (35.6%) | 25231 (37.4%) |
| **Dementia** |  |  |  |
| No | 39774 (91.9%) | 22422 (93.1%) | 62196 (92.3%) |
| Yes | 3529 (8.1%) | 1660 (6.9%) | 5189 (7.7%) |
| **Depression and/or Anxiety** |  |  |  |

|  | **Controlled BP**<br>**(N=43303)** | **Uncontrolled BP**<br>**(N=24082)** | **Overall**<br>**(N=67385)** |
| --- | --- | --- | --- |
| No | 28943 (66.8%) | 16728 (69.5%) | 45671 (67.8%) |
| Yes | 14360 (33.2%) | 7354 (30.5%) | 21714 (32.2%) |
| **Schizophrenia** |  |  |  |
| No | 42726 (98.7%) | 23853 (99.0%) | 66579 (98.8%) |
| Yes | 577 (1.3%) | 229 (1.0%) | 806 (1.2%) |

Comparisons between patients with controlled and uncontrolled BP were assessed using Pearson chi-square tests for categorical covariates and two-sample t-tests for continuous covariates. All p-values <0.001

### Multimorbidity

The overall prevalence of multimorbidity was 80.6%. Patients with multimorbidity had a higher number of annual primary care visits than patients without multimorbidity in 2017, 2018 and 2019 (**Supplementary Table S2**). As an example, 8.0% of the patients with multimorbidity had more than 9 visits in 2019 compared to 1.9% of patients without multimorbidity

### Uncontrolled blood pressure

Overall, 35.7% of patients had uncontrolled BP (34.1% in patients with multimorbidity and 42.7% in patients without multimorbidity, p<0.001). **Figure 2** presents the percentages of uncontrolled BP in patients with and without multimorbidity. Across the three age groups, patients with multimorbidity had lower rates of uncontrolled BP compared to patients without multimorbidity.

**Figure 2:**
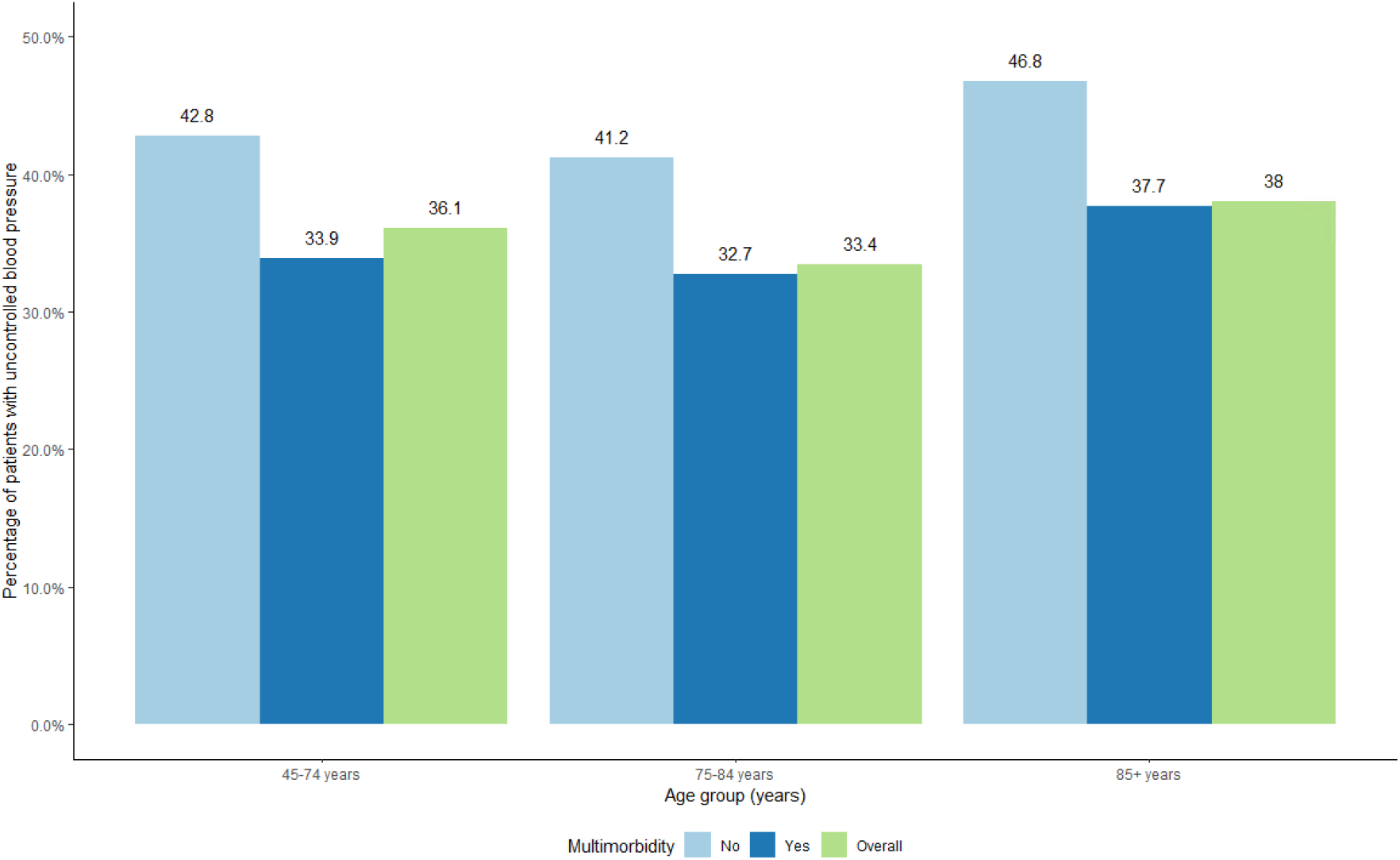
**Percentage of patients with uncontrolled blood pressure**

### The association between multimorbidity and hypertension control

Patients with multimorbidity had lower odds of uncontrolled BP compared to those without multimorbidity (adjusted OR = 0.72, 95% CI 0.68 to 0.76, adjusted for age, sex, income quintiles, smoking status, body mass index, and number of visits). Younger patients had lower odds of having uncontrolled BP than older patients. Obese patients (in class I, II and III) had higher odds of uncontrolled BP than those with normal BMI. Current smokers had higher odds of having uncontrolled BP compared to non-smokers. Increasing number of visits to primary care doctors was associated with lower odds of uncontrolled BP. (**Figure 3**)

**Figure 3:**
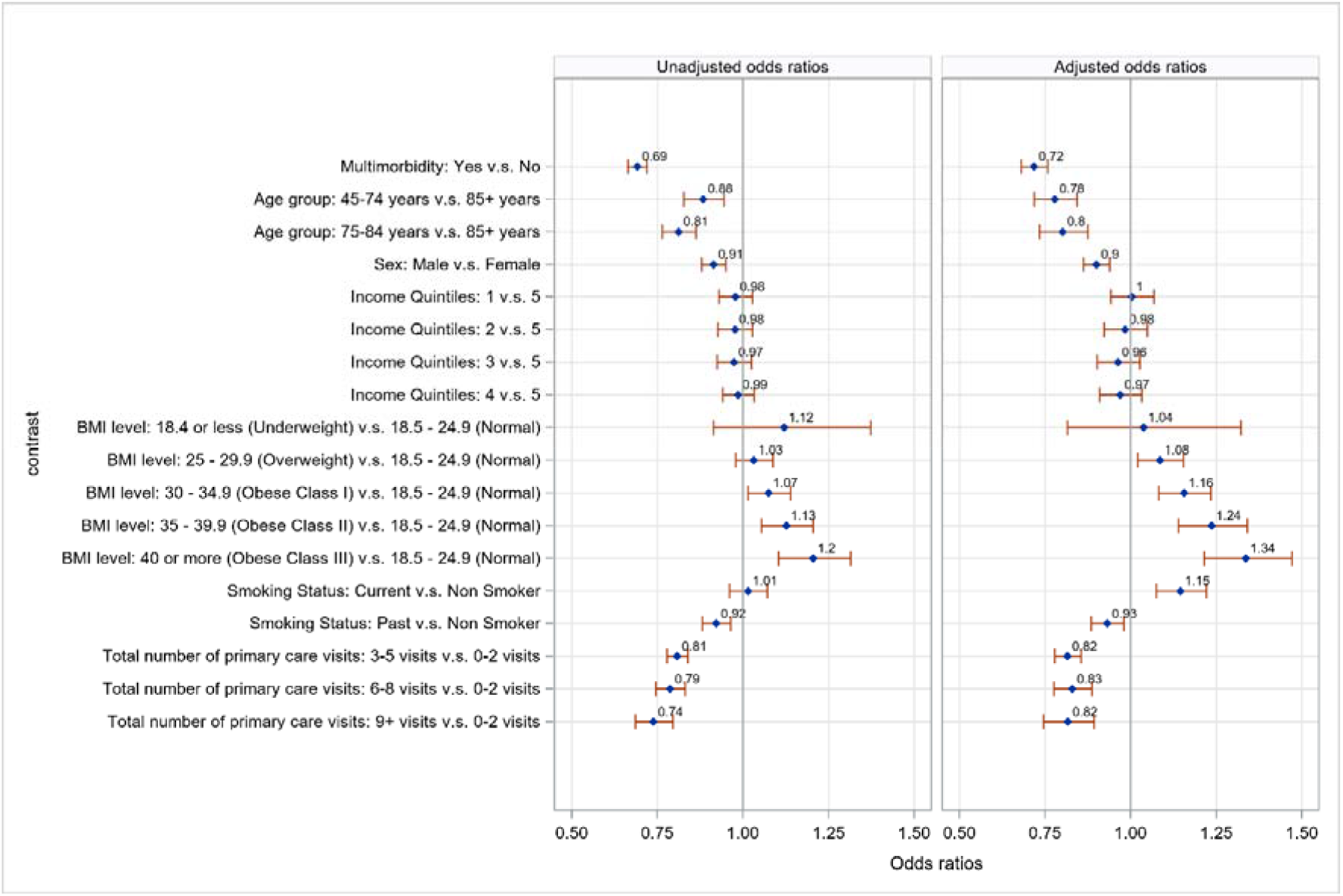
**Unadjusted and adjusted odds ratios for uncontrolled blood pressure with respect to patient characteristics. \***Odds ratio >1 means a greater likelihood of uncontrolled blood pressure. Models were adjusted for age, sex, income quintiles, smoking status, body mass index, and number of visits.

### The association between types of long-term conditions and hypertension control

Among the long-term conditions, seven conditions were significantly associated with lower likelihood of uncontrolled BP: diabetes (adjusted OR 0.73, 95%CI 0.70-0.77), heart failure (adjusted OR 0.81, 95%CI 0.73-0.91), ischemic heart disease (adjusted OR 0.74, 95%CI 0.69-0.79), schizophrenia (adjusted OR 0.79, 95%CI 0.65-0.97), depression/anxiety (adjusted OR 0.91, 95%CI 0.86-0.95), dementia (adjusted OR 0.87, 95%CI 0.80-0.95), and osteoarthritis (adjusted OR 0.89, 95%CI 0.85-0.93). The remaining long-term conditions (atrial fibrillation, COPD, asthma, chronic kidney disease, and cancer) were not associated with uncontrolled BP. (**Figure 4**)

**Figure 4:**
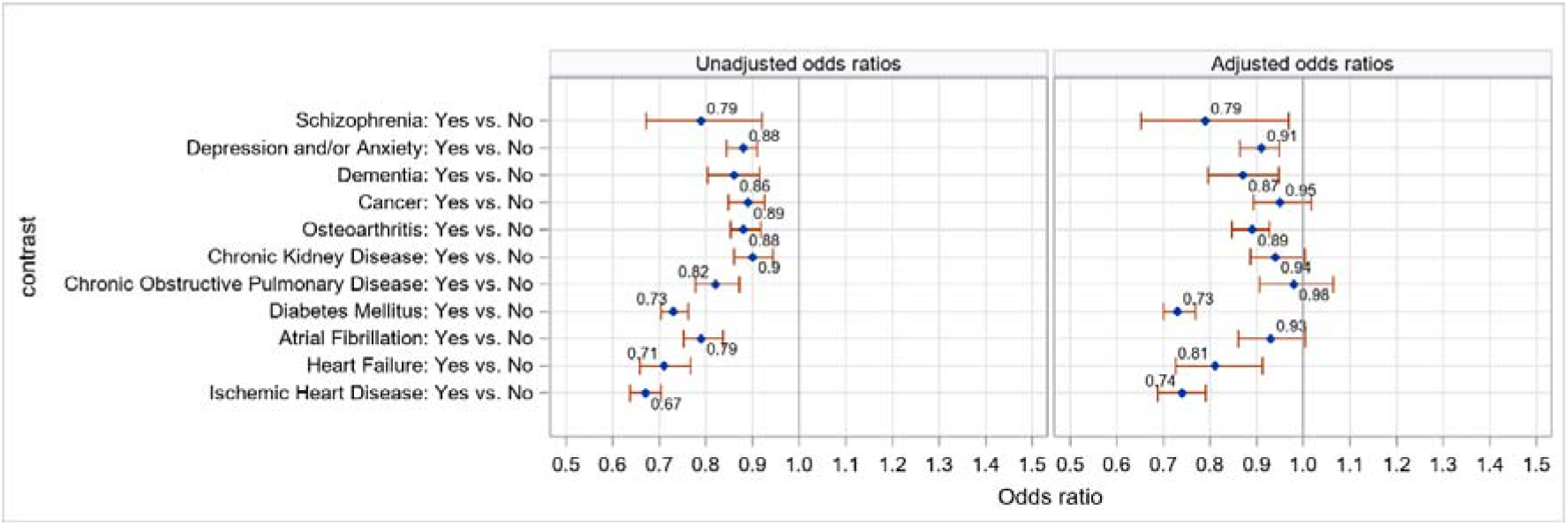
**Unadjusted and adjusted odds ratios for uncontrolled blood pressure with respect to individual long-term conditions. \***Odds ratio >1 means a greater likelihood of uncontrolled blood pressure. Models were adjusted for age, sex, income quintiles, smoking status, body mass index, and number of visits.

### Sensitivity analysis

Sensitivity analyses showed consistent results (**Supplementary Tables S3, S4**). In GEE models using ≥ 130/80 mmHg as the threshold for sub-optimal blood pressure control, multimorbidity was still significantly associated with reduced likelihoods of uncontrolled BP (adjusted ORs 0.56, 95%CI 0.54-0.59), adjusting for age, sex, income quintiles, body mass index, and number of visits).

## Discussion

### Summary of findings

In this study, we aimed to investigate the association between multimorbidity and uncontrolled BP, and to assess if individual long-term conditions are associated with uncontrolled BP. Our analysis in 67,385 Canadians aged 45 years or more with hypertension attending primary care from 2017 to 2019 showed an association between the presence of comorbidities and better blood pressure control. Multimorbidity was associated with approximately 30% lower odds of uncontrolled BP. The individual long-term conditions that were significantly associated with lower likelihood of uncontrolled BP include diabetes, heart failure, ischemic heart disease, schizophrenia, depression/anxiety, dementia, or osteoarthritis.

### Comparison to the literature

Our finding aligns with other studies that examined this association, which reported that multimorbidity was one of the strongest predictors of controlled BP in patients with hypertension. Tapela and colleagues found that multimorbidity was positively associated with BP control among participants with hypertension in the UK Biobank, and the types of comorbidities that were significantly associated with controlled BP included cardiovascular disease (OR 2.11, 95% CI 2.04 to 2.19), diabetes (OR 1.32, 95% CI 1.27 to 1.36), migraines (OR 1.68, 95% CI 1.56 to 1.81), and depression (OR 1.27, 95% CI 1.20 to 1.34).^28^ Another study conducted by Sarkar and colleagues in 31,676 patients with hypertension in the Lambeth Data-Net, a patient-level primary care database in the UK, found that 16,140 (51 %) had multimorbidity, and hypertensive patients with multimorbidity had lower BP than those with hypertension alone.^12^. Mini and colleagues also reported that blood pressure control was better in participants who reported any comorbidity (OR 2.37, 95%CI 1.51–3.71) compared to those who did not report any.^16^ However, several other studies reported the opposite findings - people with more comorbidities had poorer management and control of hypertension. In a study of 223,286 patients with hypertension in Hongkong, Wong et al. found that the proportion of patients having poor BP control increased from 35.0% to 65.0% and 69.1% when the number of medical conditions increased from zero to one and two, respectively.^15^ However, this study enrolled adult patients prescribed their first antihypertensive agents; hence people with comorbidities (such as diabetes, heart failure, coronary heart disease) who were already on a relevant medication (beta-blockers, ACE inhibitors) for these reasons would be excluded from the hypertension cohort. A cross-sectional analysis of the World Health Organisation Study of Global Ageing and Adult Health (WHO SAGE) Wave 1 (2007–10) in 41,557 adults (9778 with hypertension) from 6 middle-income countries (including China, Ghana, India, Mexico, Russia and South Africa) reported that more comorbidities were associated with increased odds of uncontrolled hypertension.^17^

The BP control rate in our study is also consistent with the literature. In a recent report from the NCD Risk Factor Collaboration using data from 1990 to 2019 on people aged 30–79 years from population-representative studies with measurement of blood pressure, the estimated rate of uncontrolled BP in Canada was 25%-30% in 2019.^29^ A report from the Danish General Practice Database of 37,651 patients with hypertension from 231 general practices showed that the overall BP control rate was 33.2%.^18^ A cross-sectional population-based study including 99,468 participants with hypertension enrolled in the UK Biobank reported that the overall control rate of BP was 38.1%.^28^

### Implication for practice and research

The results from this study may reflect the achievement of primary care in controlling cardiovascular risk factors in patients with multimorbidity, including regular BP measures and treatment. Comorbidities may be associated with more frequent healthcare utilisation and hence increase the monitoring of chronic conditions, including BP measurement and counselling on lifestyle modifications and medication adherence. Another explanation could be the study’s participation selection processes. Among people with hypertension as their only chronic health condition, those with poorer BP control may be more motivated to attend primary care and hence were included in this cohort. Thus, the observed associations could be explained by selection bias – hypertensive patients with no comorbidities, but uncontrolled BP may have been more likely to seek primary care compared to hypertensive patients with no comorbidities and controlled BP. These lead to opportunities to measure BP and obtain scripts, reminders given to them on taking medications, and better lifestyle modifications. More studies are needed to explore the mechanisms underlying the associations between multimorbidity and BP control.

### Strengths and limitations

This is one of the largest analyses of hypertension control in adults attending primary care in Canada, with comprehensive sociodemographic and high-quality detailed clinical information, including comorbidities. However, the data was only collected from patients in Ontario and is not nationally representative. Another limitation of this study is that EMR data may be incomplete, and we might be unable to capture all cases of hypertension and/or prescriptions of anti-hypertensive medications. Lifestyle modification, which is an essential strategy in the management of hypertension and can influence BP control, could not be captured through the EMR data. Duration and severity of hypertension and other chronic diseases were also not captured. Therefore, results should be replicated and cautiously interpreted before generalizing to all patients in primary care.

## Conclusion

In this large-scale study of patients attending primary care with hypertension, multimorbidity was associated with better BP control. Several long-term conditions were associated with better control, including diabetes, heart failure, ischemic heart disease, schizophrenia, depression/anxiety, dementia, and osteoarthritis. Further large-scale mixed methods studies are needed to explore the mechanisms underlying the associations between multimorbidity and BP control, and to understand barriers to BP control in primary care.

## Data Availability

The data may contain patient-related information, and therefore due to privacy concerns, has not been made publicly available. The data that was used in this study is available from UTOPIAN at the Department of Family and Community Medicine, Temerty Faculty of Medicine, University of Toronto, following Research Ethics Board approval; please contact

## Acknowledgements

Dr Tu Nguyen is supported by the University of Sydney Global Development Awards.

## Supplementary data

**Figure S1:**
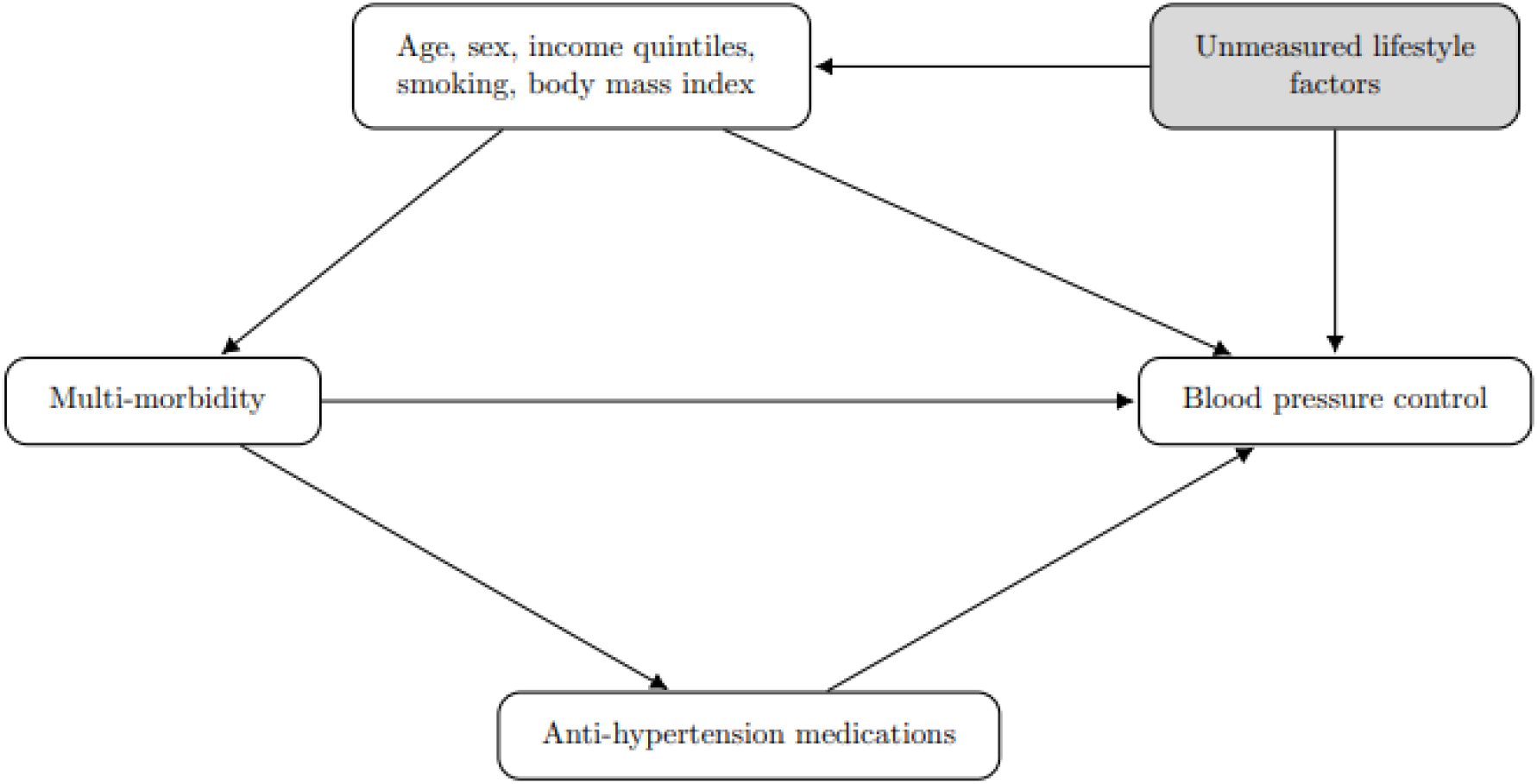
Directed acyclic graph for the effect of multi-morbidity on blood pressure control.

**Table S1:**
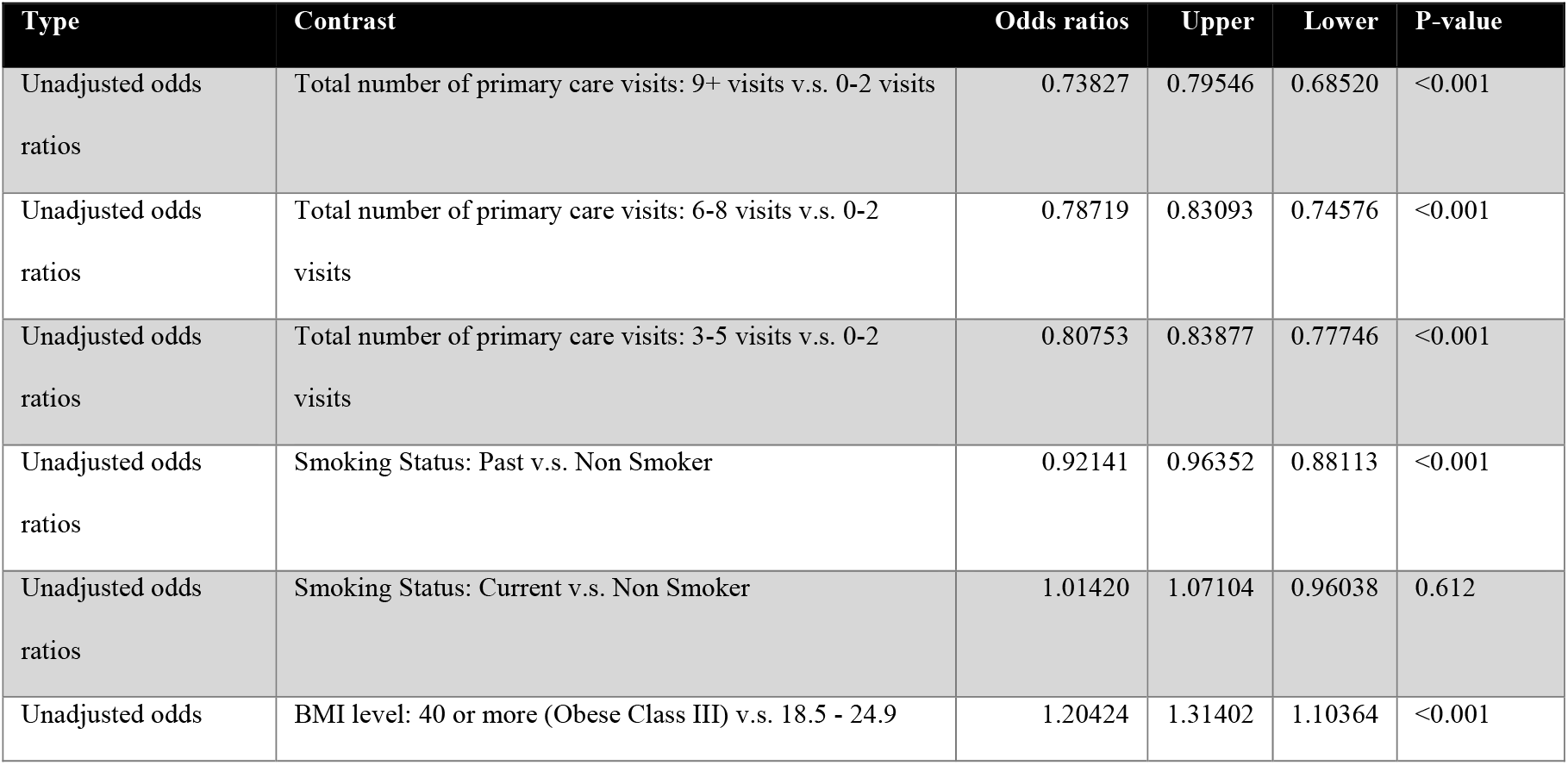
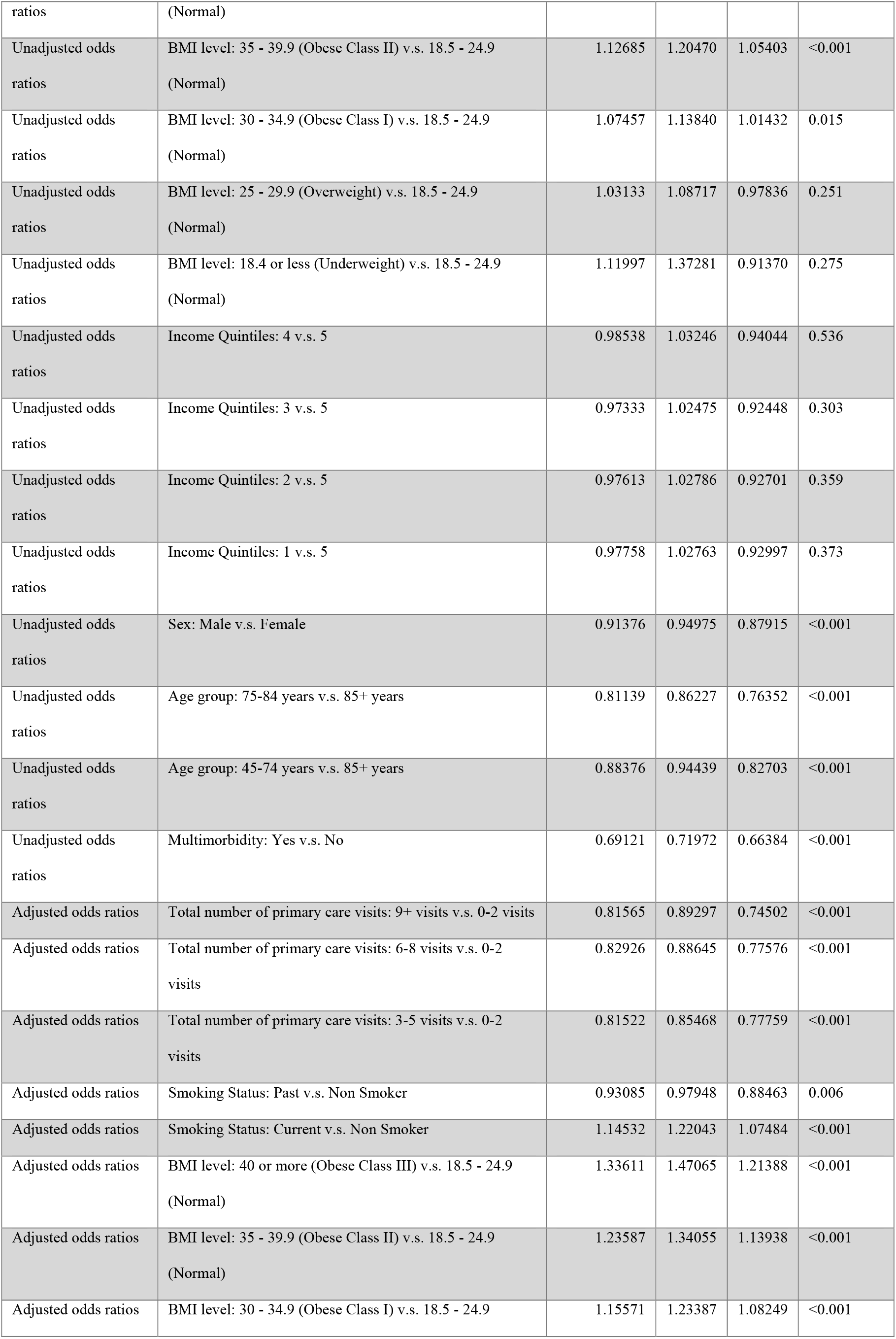
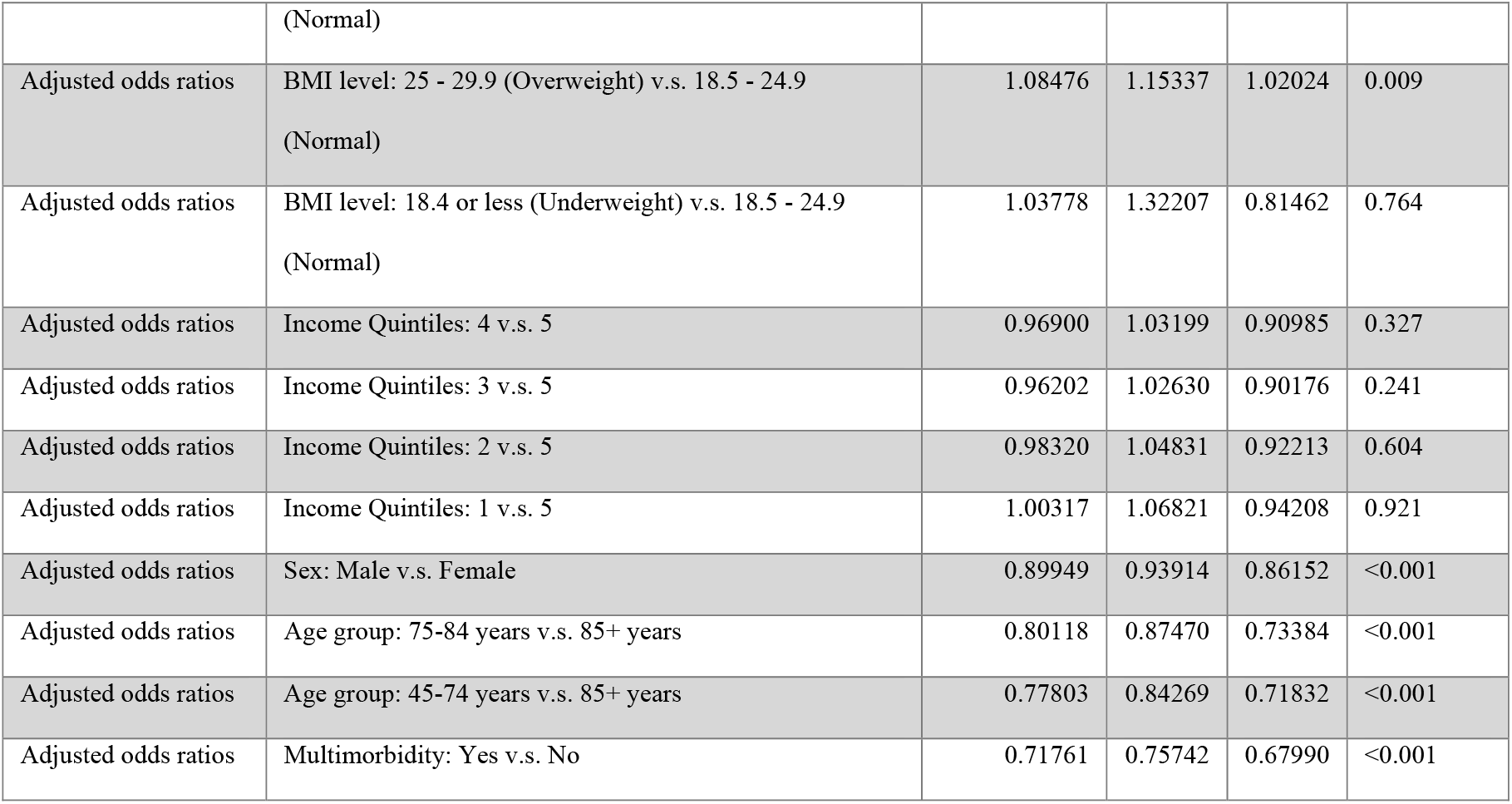
Unadjusted and adjusted odds ratios for uncontrolled blood pressure with respect to patient characteristics.

**Table S2:**
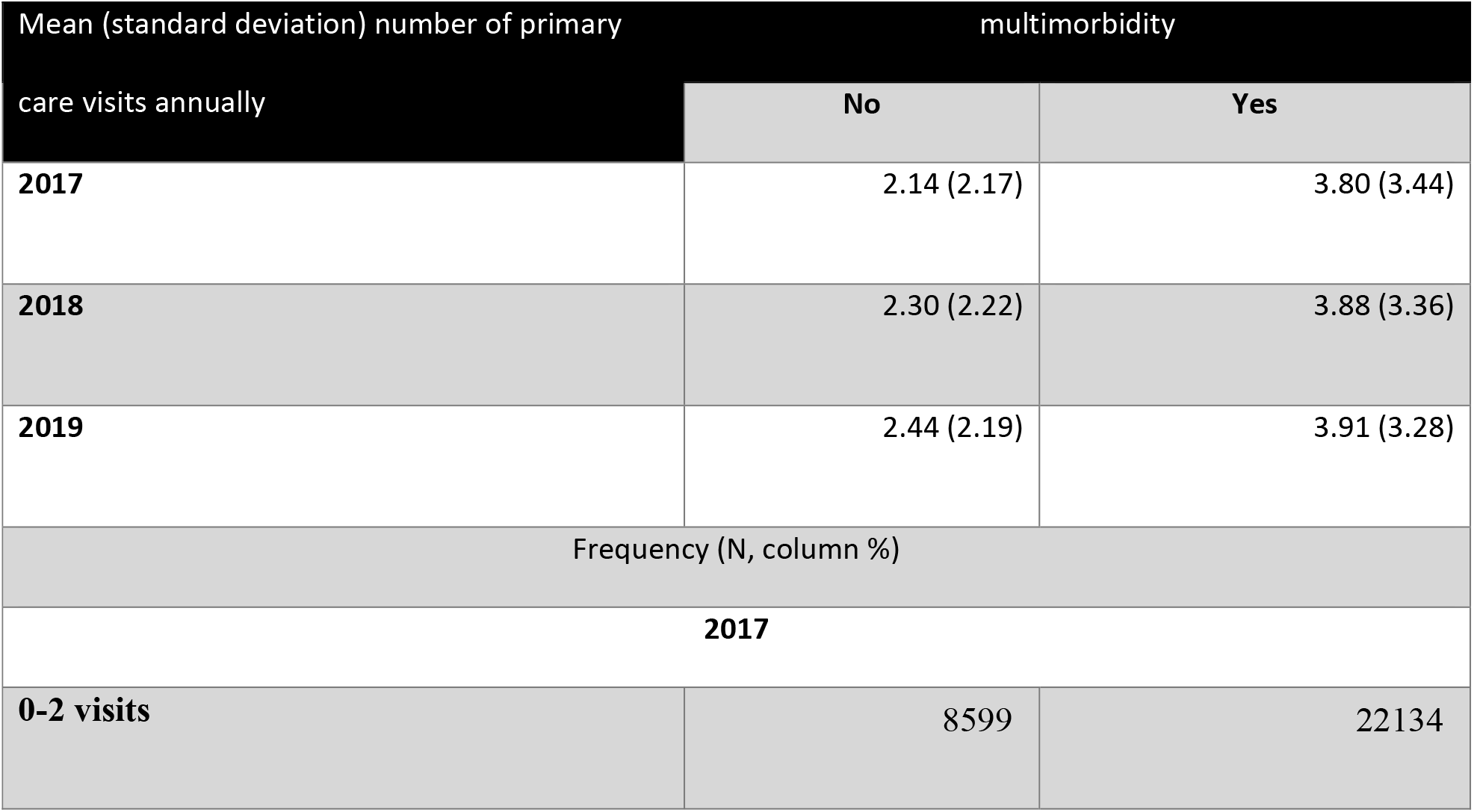
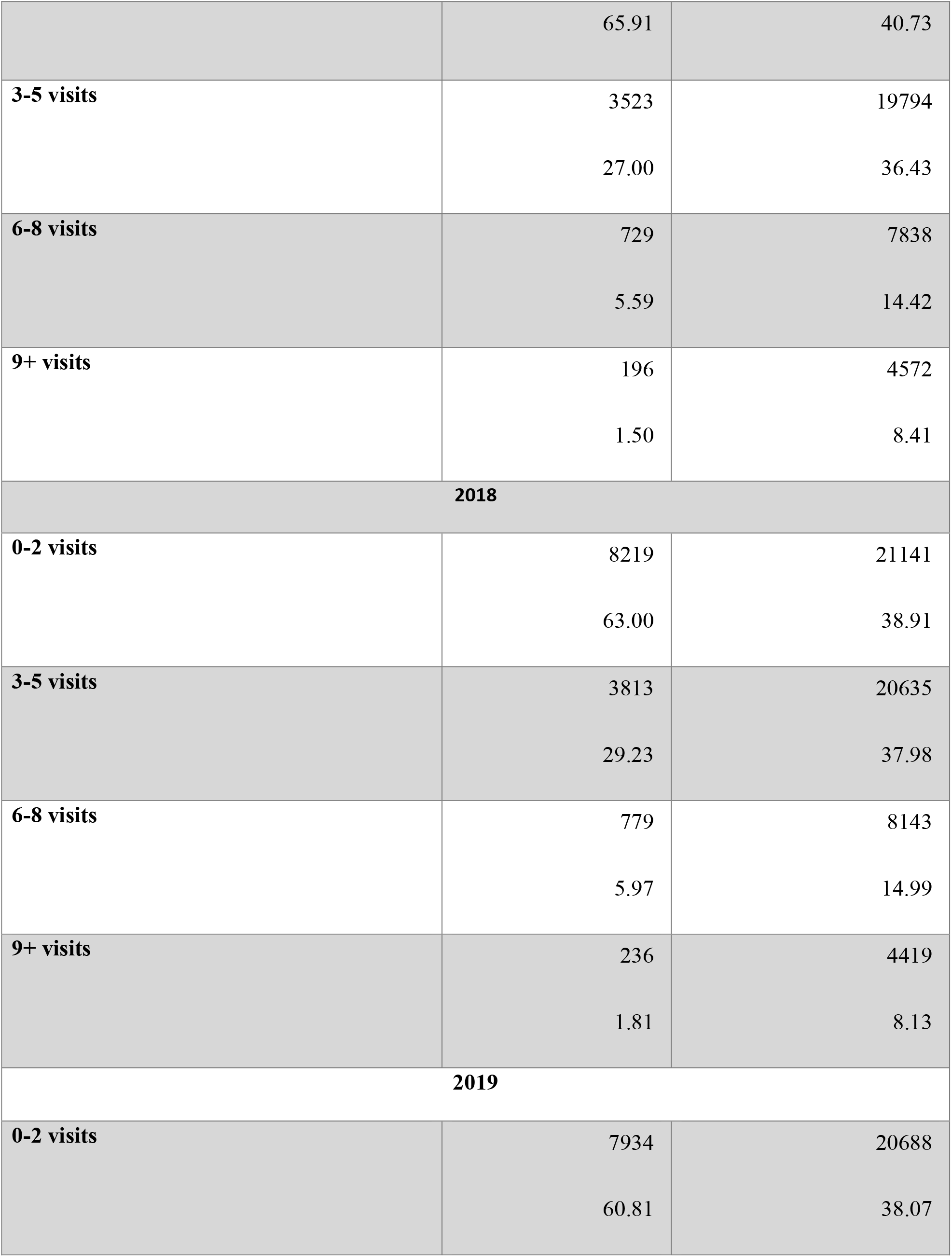
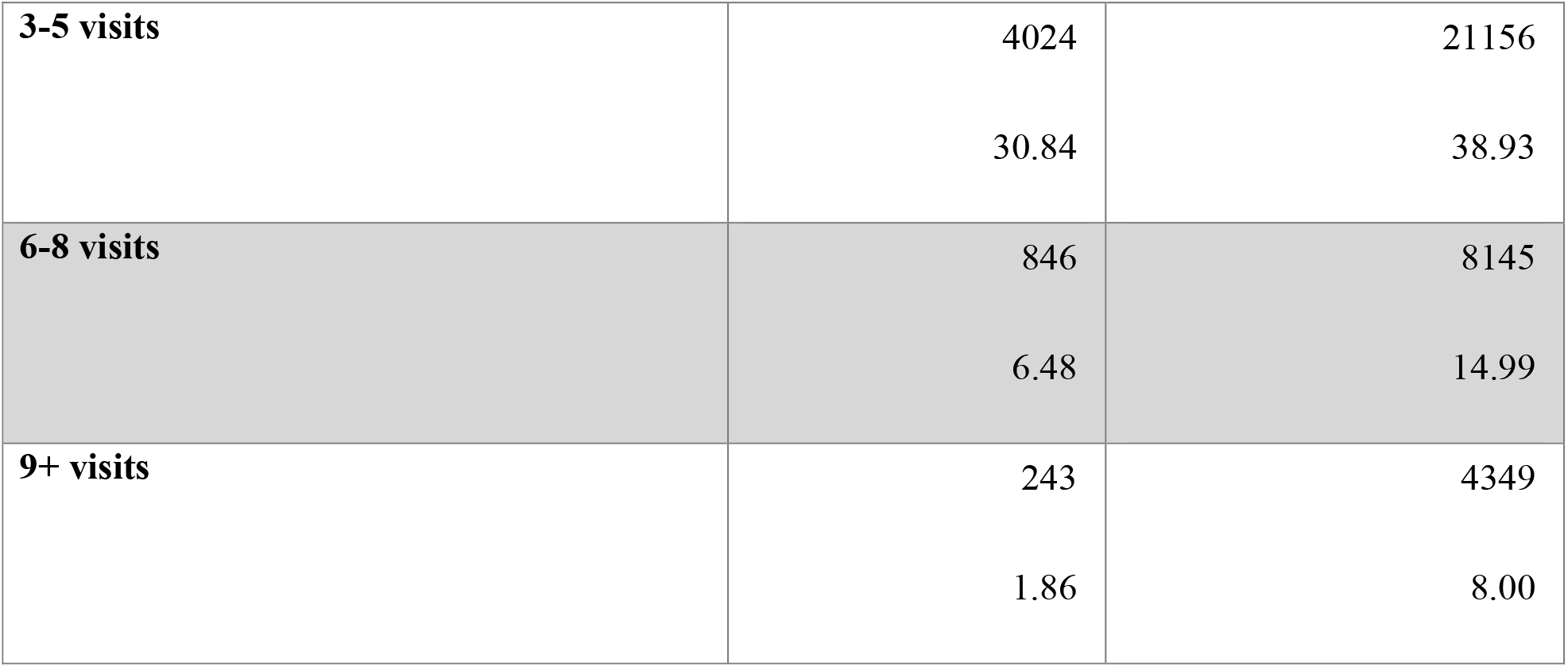
Mean number of primary care visits for patients with and without multi-morbidity (annually)

| Mean (standard deviation) number of primary care visits annually |  | multimorbidity |
| --- | --- | --- |
|  | No | Yes |
| **2017** | 2.14 (2.17) | 3.80 (3.44) |
| **2018** | 2.30 (2.22) | 3.88 (3.36) |
| **2019** | 2.44 (2.19) | 3.91 (3.28) |
| Frequency (N, column %) |  |  |
| **2017** |  |  |
| **0-2 visits** | 8599 | 22134 |
|  | 65.91 | 40.73 |
| **3-5 visits** | 3523 | 19794 |
|  | 27.00 | 36.43 |
| **6-8 visits** | 729 | 7838 |
|  | 5.59 | 14.42 |
| **9+ visits** | 196 | 4572 |
|  | 1.50 | 8.41 |
| **2018** |  |  |
| **0-2 visits** | 8219 | 21141 |
|  | 63.00 | 38.91 |
| **3-5 visits** | 3813 | 20635 |
|  | 29.23 | 37.98 |
| **6-8 visits** | 779 | 8143 |
|  | 5.97 | 14.99 |
| **9+ visits** | 236 | 4419 |
|  | 1.81 | 8.13 |
| **2019** |  |  |
| **0-2 visits** | 7934 | 20688 |
|  | 60.81 | 38.07 |
| **3-5 visits** | 4024 | 21156 |
|  | 30.84 | 38.93 |
| **6-8 visits** | 846 | 8145 |
|  | 6.48 | 14.99 |
| **9+ visits** | 243 | 4349 |
|  | 1.86 | 8.00 |

**Table S3:** Patient characteristics for sub-optimal blood pressure control (≥130/80 mmHg)

|  | Controlled BP<br>(N=19535) | Uncontrolled BP<br>(N=47850) | Overall<br>(N=67385) | P-value |
| --- | --- | --- | --- | --- |
| **Age (years)** |  |  |  |  |
| Mean (SD) | 72.1 (11.5) | 69.2 (11.9) | 70.1 (11.8) | <0.001 |
| Median [Min, Max] | 72.0 [47.0, 111] | 69.0 [47.0, 110] | 70.0 [47.0, 111] |  |
| **Age group (years)** |  |  |  |  |
| 45-74 years | 12809 (65.6%) | 35009 (73.2%) | 47818 (71.0%) | <0.001 |
| 75-84 years | 4597 (23.5%) | 8816 (18.4%) | 13413 (19.9%) |  |
| 85+ years | 2129 (10.9%) | 4025 (8.4%) | 6154 (9.1%) |  |
| **Sex** |  |  |  |  |
| Female | 10183 (52.1%) | 25586 (53.5%) | 35769 (53.1%) | 0.00652 |
| Male | 9352 (47.9%) | 22264 (46.5%) | 31616 (46.9%) |  |
| **Income Quintiles** |  |  |  |  |
| 1 (low income) | 4259 (21.8%) | 9815 (20.5%) | 14074 (20.9%) | <0.001 |
| 2 | 3605 (18.5%) | 8535 (17.8%) | 12140 (18.0%) |  |
| 3 | 3223 (16.5%) | 7979 (16.7%) | 11202 (16.6%) |  |
| 4 | 3323 (17.0%) | 8426 (17.6%) | 11749 (17.4%) |  |
| 5 (high income) | 4626 (23.7%) | 12043 (25.2%) | 16669 (24.7%) |  |
| Missing | 499 (2.6%) | 1052 (2.2%) | 1551 (2.3%) |  |
| Multimorbidity |  |  |  |  |
| No | 2528 (12.9%) | 10519 (22.0%) | 13047 (19.4%) | <0.001 |
| Yes | 17007 (87.1%) | 37331 (78.0%) | 54338 (80.6%) |  |
| Body mass index level |  |  |  |  |
| Missing | 3347 (17.1%) | 9502 (19.9%) | 12849 (19.1%) | <0.001 |
| 18.4 or less (Underweight) | 152 (0.8%) | 260 (0.5%) | 412 (0.6%) |  |
| 18.5 - 24.9 (Normal) | 3226 (16.5%) | 6447 (13.5%) | 9673 (14.4%) |  |
| 25 - 29.9 (Overweight) | 6190 (31.7%) | 14279 (29.8%) | 20469 (30.4%) |  |
| 30 - 34.9 (Obese Class I) | 4043 (20.7%) | 10330 (21.6%) | 14373 (21.3%) |  |
| 35 - 39.9 (Obese Class II) | 1630 (8.3%) | 4360 (9.1%) | 5990 (8.9%) |  |
| 40 or more (Obese Class III) | 947 (4.8%) | 2672 (5.6%) | 3619 (5.4%) |  |
| Most recent smoking status |  |  |  |  |
| Missing | 4935 (25.3%) | 12867 (26.9%) | 17802 (26.4%) | <0.001 |
| Current | 1909 (9.8%) | 4852 (10.1%) | 6761 (10.0%) |  |
| Non Smoker | 8856 (45.3%) | 22011 (46.0%) | 30867 (45.8%) |  |
| Past | 3835 (19.6%) | 8120 (17.0%) | 11955 (17.7%) |  |
| Cancer |  |  |  |  |
| No | 16955 (86.8%) | 42275 (88.3%) | 59230 (87.9%) | <0.001 |
| Yes | 2580 (13.2%) | 5575 (11.7%) | 8155 (12.1%) |  |
| Ischemic Heart disease |  |  |  |  |
| No | 16032 (82.1%) | 42829 (89.5%) | 58861 (87.4%) | <0.001 |
| Yes | 3503 (17.9%) | 5021 (10.5%) | 8524 (12.6%) |  |
| Heart failure |  |  |  |  |
| No | 18006 (92.2%) | 45830 (95.8%) | 63836 (94.7%) | <0.001 |
| Yes | 1529 (7.8%) | 2020 (4.2%) | 3549 (5.3%) |  |
| Atrial fibrillation |  |  |  |  |
| No | 16799 (86.0%) | 43441 (90.8%) | 60240 (89.4%) | <0.001 |
| Yes | 2736 (14.0%) | 4409 (9.2%) | 7145 (10.6%) |  |
| Diabetes Mellitus |  |  |  |  |
| No | 11842 (60.6%) | 33494 (70.0%) | 45336 (67.3%) | <0.001 |
| Yes | 7693 (39.4%) | 14356 (30.0%) | 22049 (32.7%) |  |
| Chronic Kidney disease |  |  |  |  |
| No | 15193 (77.8%) | 40120 (83.8%) | 55313 (82.1%) | <0.001 |
| Yes | 4342 (22.2%) | 7730 (16.2%) | 12072 (17.9%) |  |
| Chronic Obstructive Pulmonary Disease |  |  |  |  |
| No | 17487 (89.5%) | 44075 (92.1%) | 61562 (91.4%) | <0.001 |
| Yes | 2048 (10.5%) | 3775 (7.9%) | 5823 (8.6%) |  |
| Mental health |  |  |  |  |
| No | 11804 (60.4%) | 30997 (64.8%) | 42801 (63.5%) | <0.001 |
| Yes | 7731 (39.6%) | 16853 (35.2%) | 24584 (36.5%) |  |

**Table S4:** Adjusted odds ratios using GEE model for sensitivity analysis using ≥ 130/80 mmHg as the threshold for sub-optimal blood pressure control.

| Contrast | Adjusted odds ratios | Lower | Upper | P-value |
| --- | --- | --- | --- | --- |
| Multimorbidity: Yes vs. No | 0.56 | 0.54 | 0.59 | <0.001 |
| Age group: 45-74 years vs. 85+ years | 1.29 | 1.22 | 1.37 | <0.001 |
| Age group: 75-84 years vs. 85+ years | 0.98 | 0.92 | 1.05 | 0.582 |
| Sex: Male vs. Female | 0.91 | 0.88 | 0.94 | <0.001 |
| Income Quintiles: 1 vs. 5 | 0.93 | 0.88 | 0.98 | 0.005 |
| Income Quintiles: 2 vs. 5 | 0.95 | 0.90 | 1.00 | 0.062 |
| Income Quintiles: 3 vs. 5 | 0.97 | 0.92 | 1.02 | 0.34 |
| Income Quintiles: 4 vs. 5 | 0.98 | 0.93 | 1.03 | 0.405 |
| Region: Urban vs. Rural | 0.84 | 0.78 | 0.90 | <0.001 |

**Table S5:**
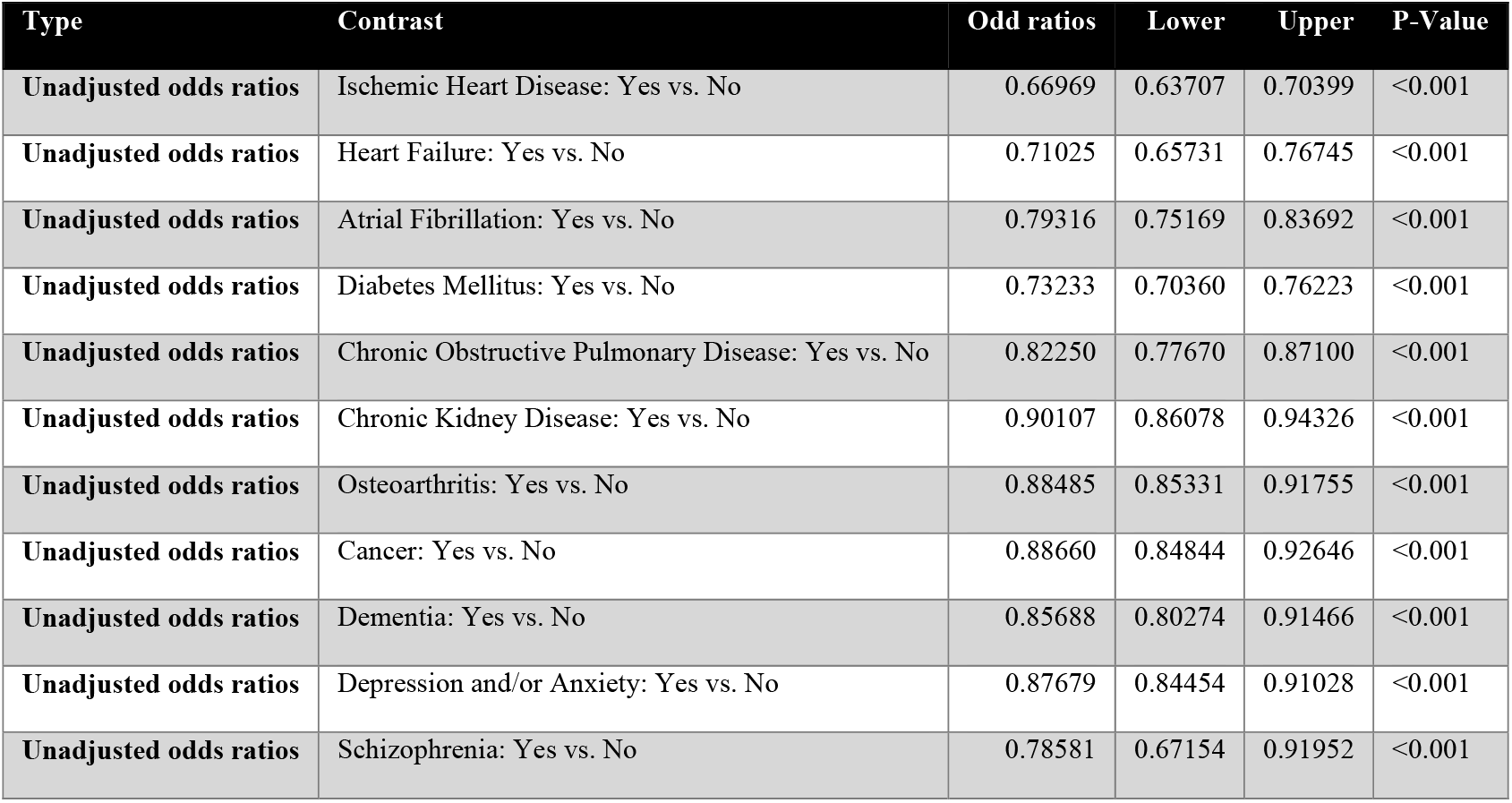
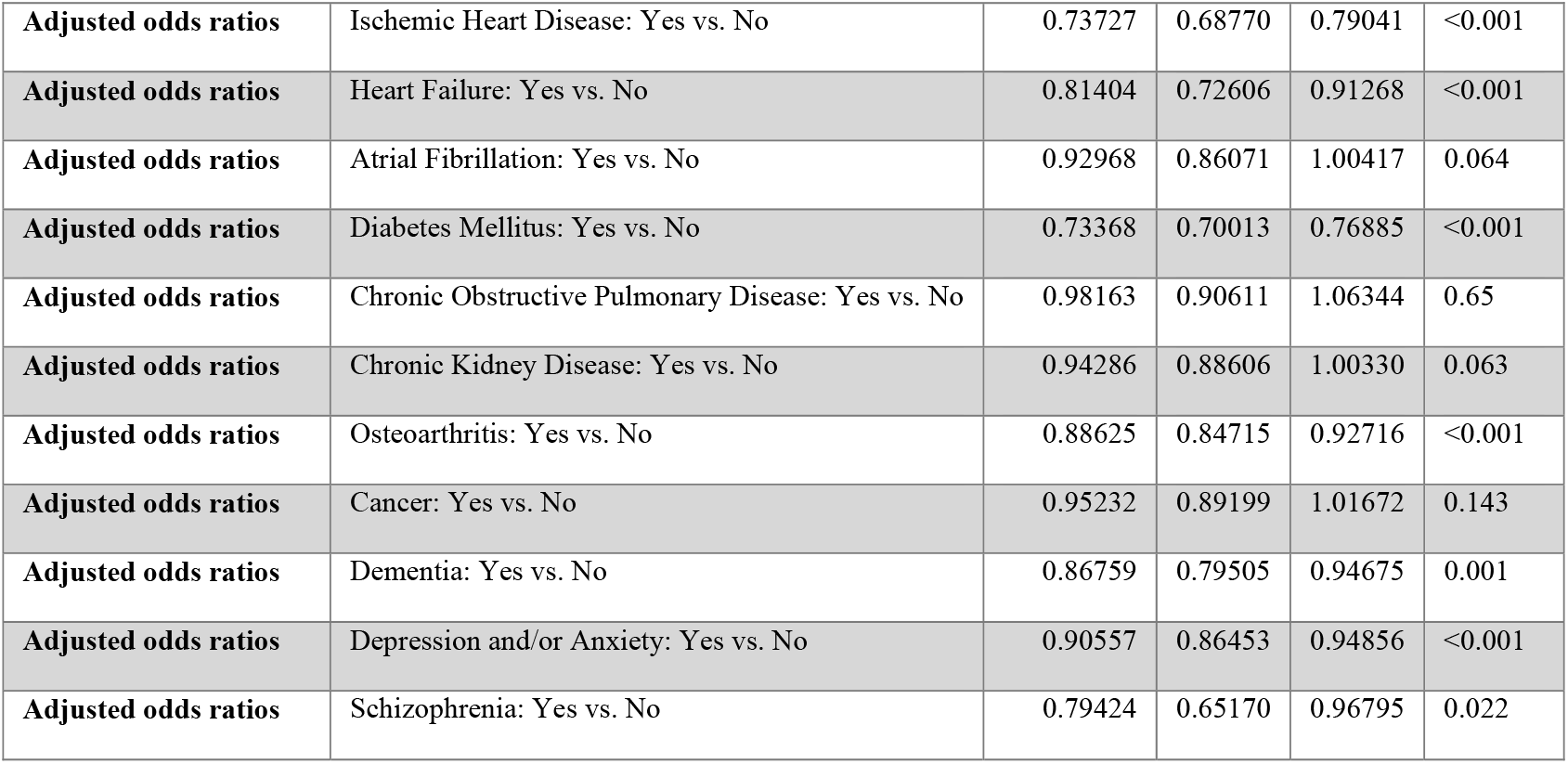
Unadjusted and adjusted odds ratios for uncontrolled blood pressure with respect to co-morbidities.

### Appendix

The appendix section contains the following sections:

i. Drug names for different classes of hypertension medications;
ii. Definition for the hypertension phenotype
iii. Definition for primary care visit in UTOPIAN database

### Hypertension medication classes

The following search criteria were used to identify hypertension medication:

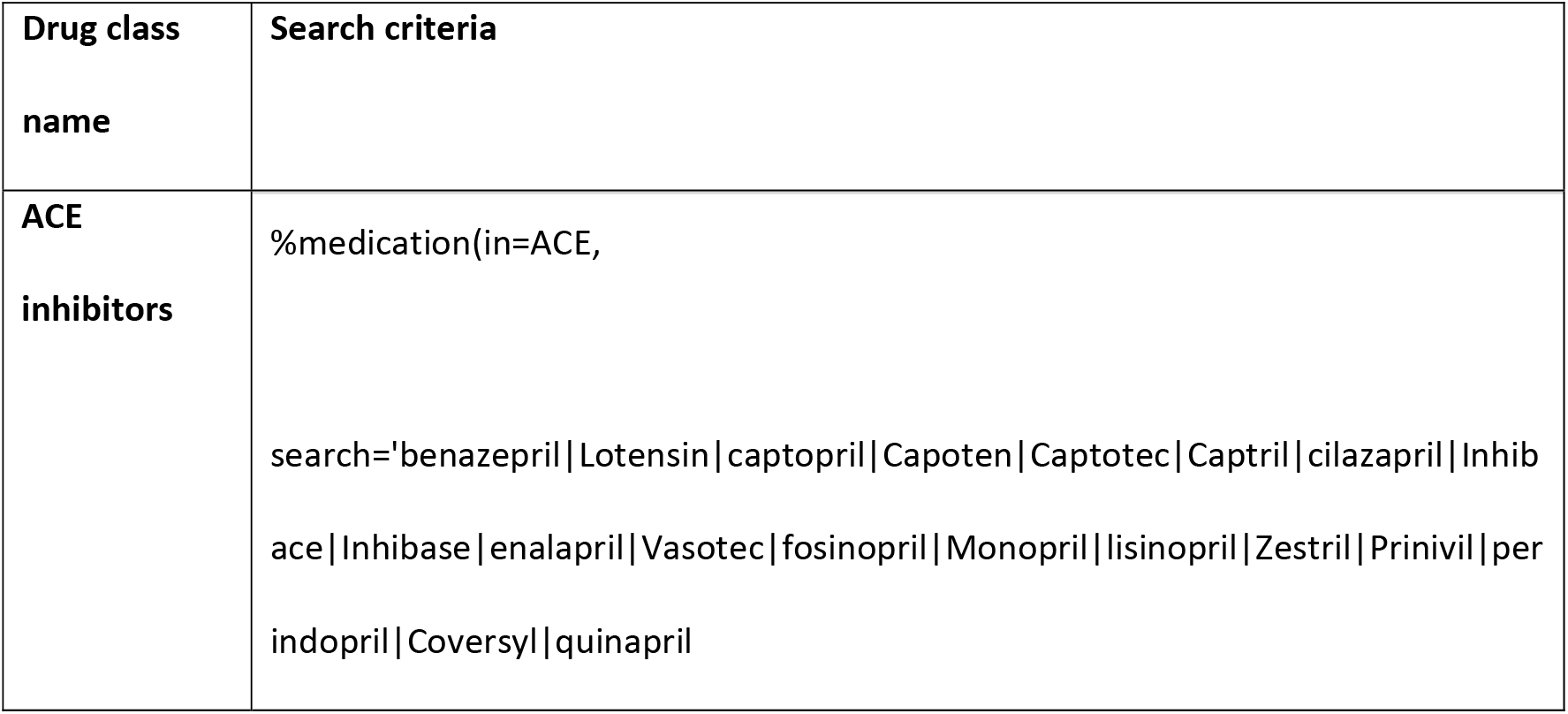
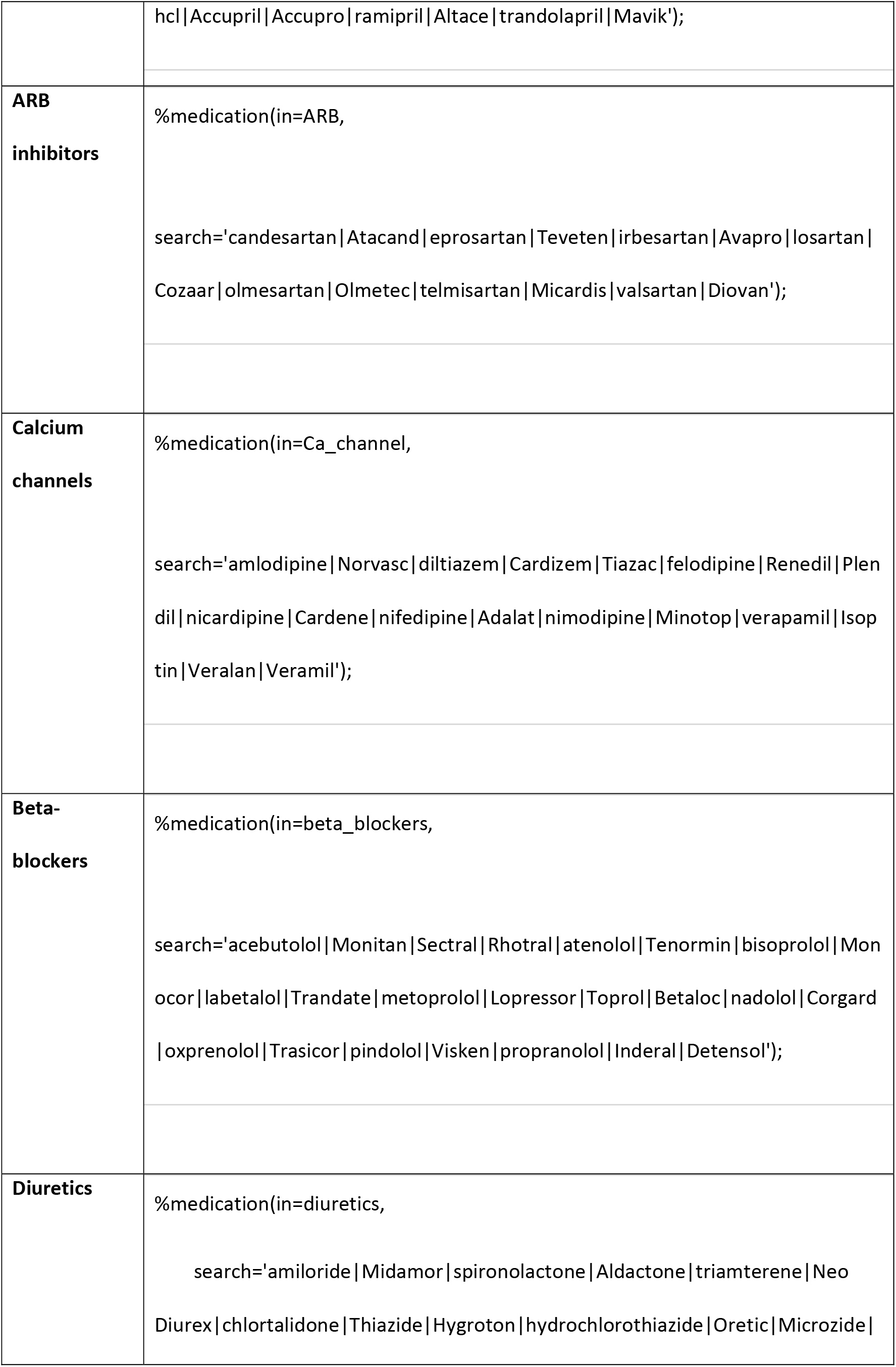
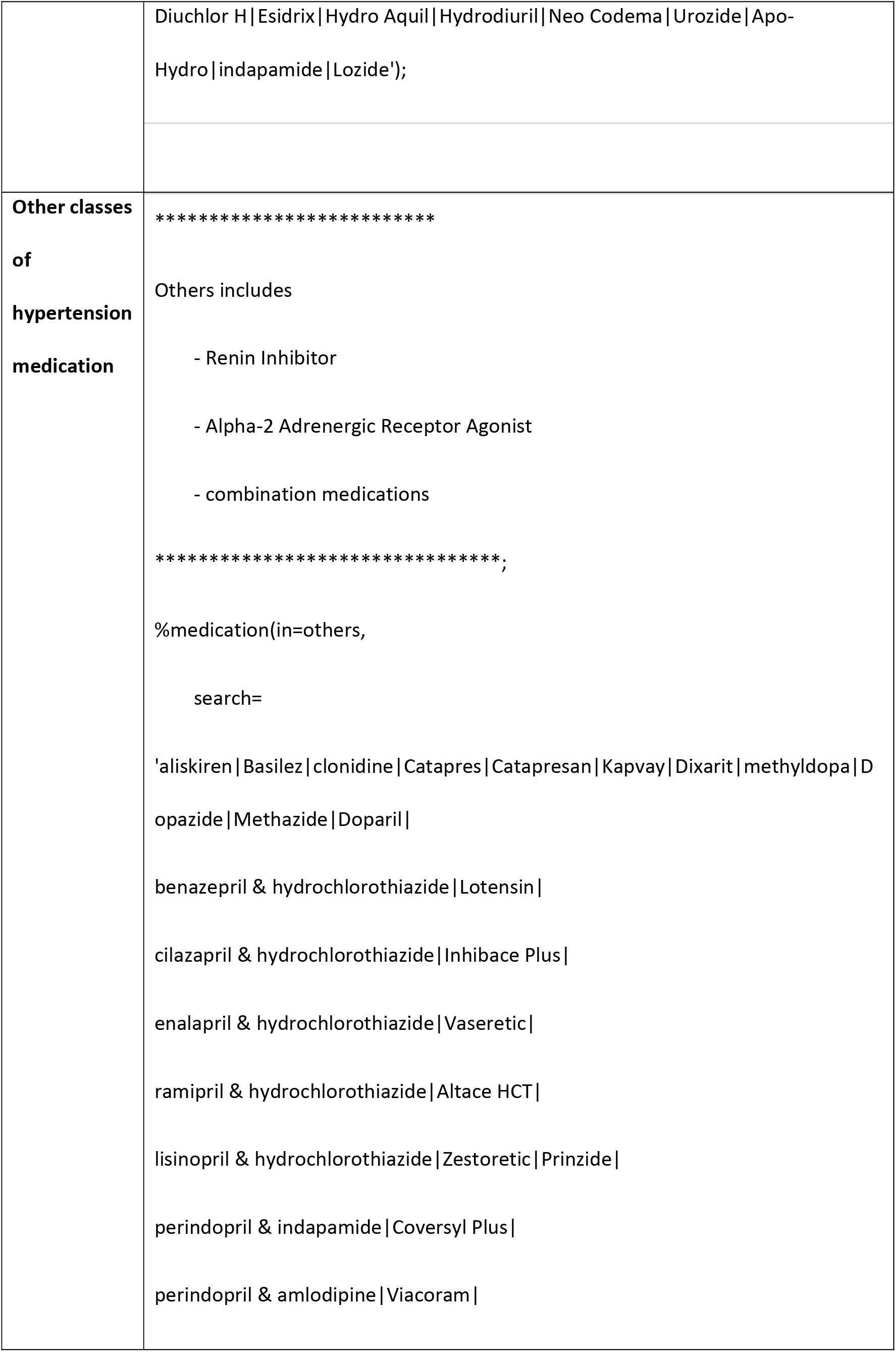
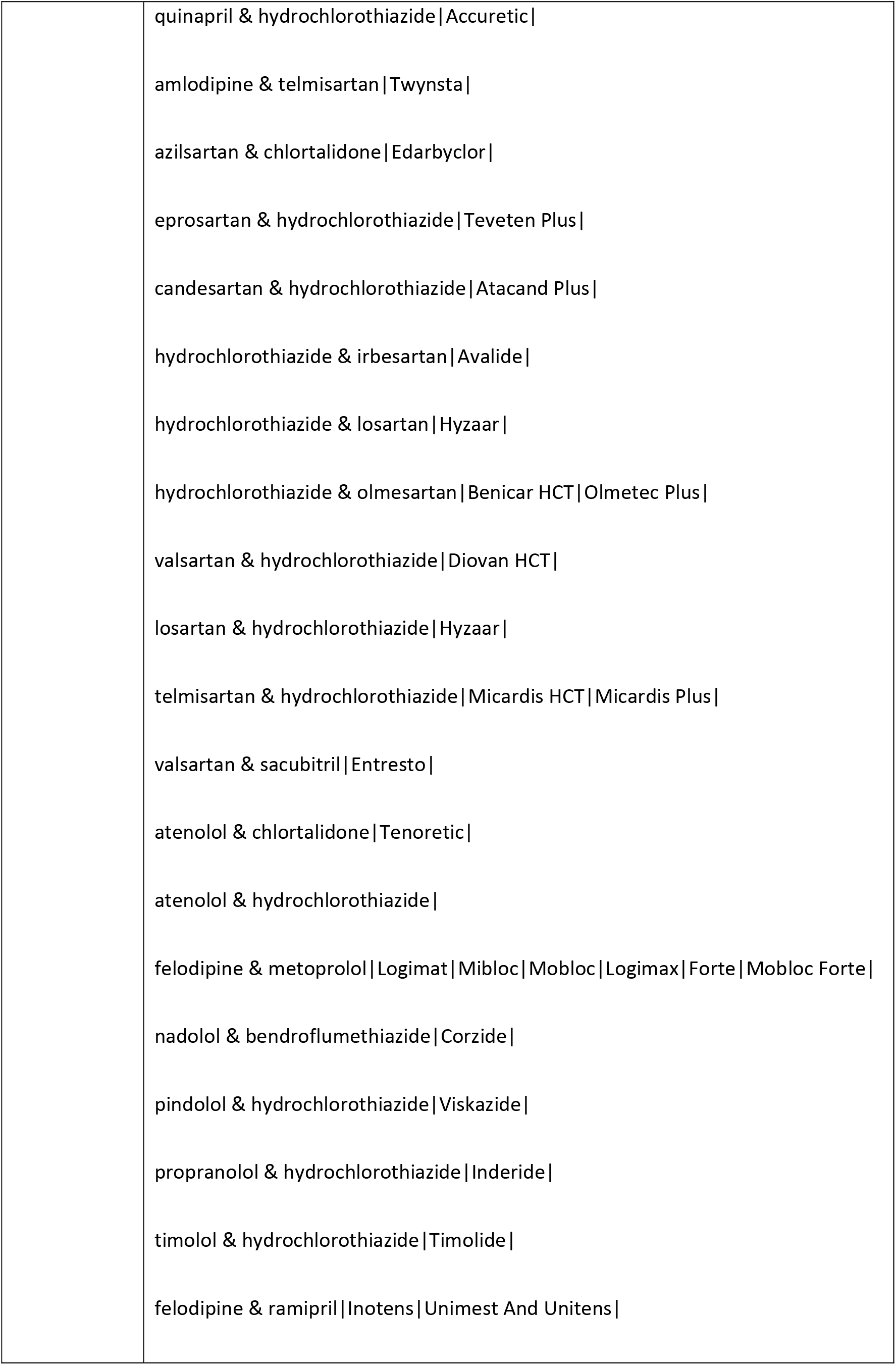
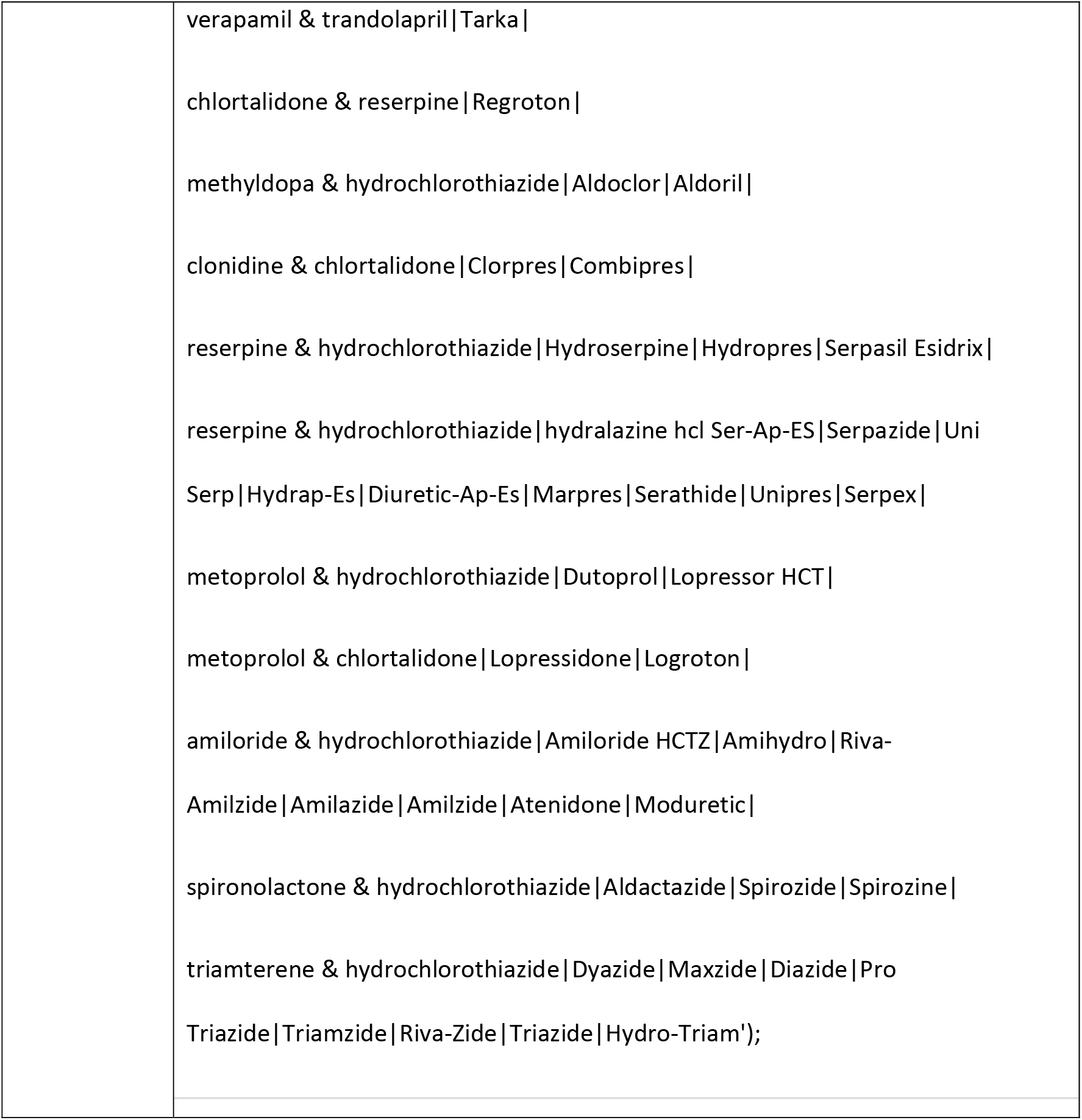

### Hypertension phenotype

We defined the hypertension phenotype using the following criteria:

1. Free text documentation of hypertension in the past or present health condition section of the cumulative patient included at least one of the following terms:

2. Anti-hypertensive medication (listed above) was prescribed and an elevated blood pressure reading was recorded at any point in the EMR

|  |  |  |
| --- | --- | --- |
| 488 | Include | Do not include |
| 489 | Hypertension | Hypertensive response |
| 490 | Hypertensive | Borderline |
| 491 | Htn | Occular/ocular |
| 492 |  | Maternal |
| 493 |  | Gestation |
| 494 |  | White coat |
| 495 |  | Pulmonary |
| 496 |  | Pregnancy induced/PIH |

a. elevated blood pressure reading is defined as systolic blood pressure >= 140 mmHg or diastolic blood pressure >= 90 mmHg OR
3. Anti-hypertensive medication (listed above) was prescribed and a billing record with the diagnosis code for hypertension (401) was found at any point in the EMR OR
4. A billing record with the diagnosis code for hypertension (401) was found and an elevated blood pressure reading was recorded at any point in the EMR

a. elevated blood pressure reading is defined as systolic blood pressure >= 140 mmHg or diastolic blood pressure >= 90 mmHg

### Definition for primary care visit in UTOPIAN database

OHIP service codes billed during the study period (Jan 2017 to Dec 2019) were used to select family physician visits that occurred via telephone, video, or in-person. Billing records for eligible patients containing any of the following service codes were counted as family physician visits:

| Code | Description | Code | Description |
| --- | --- | --- | --- |
| A001 | minor assessment | K017 | periodic health visit-child aft. 2nd birthday |
| A002 | enhanced 18-month well baby visit | K022 | hiv prim care individ care 1/2 hr or major part |
| A003 | major assessment | K028 | sexually transmitted disease (std) counseling |
| A004 | general re-assessment | K030 | diabetic management fee |
| A007 | intermediate assessment | K032 | gp-specific neurocognitive assessment |
| A008 | mini assessment | K033 | counselling - 1 pt/yr/unit |
| A071 | complex medical specific re-assessment | K039 | smoking cessation follow-up visit |
| A131 | complex medical specific re-assessment | K130 | periodic health visit - adolescent |
| A134 | medical specific re-assessment | K131 | periodic health visit - adult aged 18 to 64 inclusive |
| A624 | medical specific re-assessment | K132 | periodic health visit - adult 65 years of age and older |
| A888 | partial assessment | K680 | substance abuse - extended assessment |
| A903 | pre-op assessment | P003 | obs.-prenatal care-gen.assess-major prenatal visit |
| A920 | medical management of early pregnancy, initial visit | P004 | obs.-prenatal care-minor prenatal assess.-subseq.prenat.vis. |
| K005 | primary mental health | P005 | antenatal health screen |
| K007 | ind. psychotherapy per half hour - gp | P008 | obs.-post-natal care in office |
| K013 | counselling-one or more people-per 1/2hr | K037 | fibromyalgia/chronic fatigue syndrome care |
| K080 | Minor assessment of patient by telephone or video | K081 | Intermediate assessment including psychotherapy by telephone or video |
| K082 | Psychotherapy, psychiatric or mental health counselling by telephone or video | K087 | Minor assessment of an uninsured by telephone or video |
| K088 | Intermediate assessment of an uninsured patient including psychotherapy by telephone or video | K089 | Psychotherapy, psychiatric or mental health counselling of an uninsured patient by telephone or video |

